# A Novel Three-way fusion image segmentation for early esophageal cancer detection

**DOI:** 10.1101/2022.08.05.22277711

**Authors:** Xintong Ren, Zhen Zhang, Junchao Jiang, Guodong Li, Jiahua Jiang, Wuwei Ren, Xinyong Jia

**Author notes:** Correspondence to Xinyong Jia, Junchao Jiang and Guodong Li,. XR, ZZ contributed equally.

## Abstract

**Objective:** Esophageal cancer (EC) is a prevalent malignancy worldwide. Early-stage esophageal cancer (EEC) diagnostics is crucial for improving patient survival. However, EC is highly aggressive with a poor prognosis, even for experienced endoscopists. To address these problems, this study aims to develop a novel computer-aided diagnosis (CAD) method to improve the accuracy and efficiency of EEC diagnostics.

**Methods:** Three-way fusion CAD method that employs multiple frameworks, including the hybrid task cascade ResNeXt101 with deformable convolutional networks, to accurately detect EC. Our method incorporates dual annotation categories on ME-NBI imaging from a local perspective and one category on LCE imaging from an broader perspective. This integration provides a substantial improvement of accuracy over traditional CAD technologies.

**Results:** Our three-way fusion CAD method achieved top performances of 0.923 mAP on ME-NBI and 0.862 mAP on LCE, demonstrating superior diagnostic performance compared to traditional CAD methods. Furthermore, the treatment boundary mAP is expected to be even higher by definition in clinical settings. Our method also achieved promising precision and recall rates of 93.98% and 93.05% for ME-NBI, and 82.89% and 88.32% for LCE, respectively.

**Conclusions:** Our novel three-way fusion CAD method accurately detects EC in both ME-NBI and LCE imaging, providing accurate treatment boundaries on both image and patient levels. Our approach shows potential for clinical application, with promising mAP, precision, and recall rates. Further work will focus on collecting and analyzing patient data to improve the method’s real-time performance in clinical settings.

## Introduction

Esophageal cancer (EC) is a significant global health challenge, ranking as the sixth highest cause of cancer-related deaths. In particular, China experiences a high incidence of EC, with nearly half of all new cases reported. Therefore, early detection and intervention are crucial for improving patient outcomes. The five-year survival rate for EC exceeds 80% [1, 2] when diagnosed at early stages. However, it drops to just 20% with late detection [3]. To improve survival rates, there is a need for effective techniques that enable earlier diagnosis and curative treatment, as well as proper screening and early detection practices.

Accurately defining the boundaries of EC is a critical challenge in clinical practice to improve the survival rate of early-stage patients. The treatment boundary is a fundamental reference for endoscopic submucosal dissection (ESD) surgery, which is essential for subsequent ESD treatment. However, even experienced endoscopists may struggle to identify early-stage EC, particularly when distinguishing intramucosal neoplasias that appear as small erosions, flat mucous membranes, or normal mucosa in biopsy specimens. Incorrect boundary definition can lead to adverse outcomes, such as scar stenosis and difficulty in eating after surgery if the boundary is larger than the actual lesion. On the other hand, if the boundary is smaller than the actual lesion, it can lead to residual lesions prone to recurrence with a poor prognosis. To address this clinical challenge, it is imperative for physicians to take a comprehensive approach that investigates both local and overall boundaries, in addition to pathological comparison.

Computer-aided diagnosis (CAD) is becoming increasingly important as a tool to improve accuracy, conserve time, lessen workload, and standardize diagnosis in EC [4]. However, traditional CAD techniques have limitations in providing accurate treatment boundaries in clinical settings, as they only simply consider the lesion boundary as a whole and do not account for the unevenness of the lesion, leading to indistinct side planes and ambiguous boundaries. Moreover, factors such as focus, shadows, and blurriness can interfere with determining the treatment boundary, making the task of precise boundary delineation even more challenging.

To overcome these challenges, we propose a novel three-way fusion CAD technology that augments the identification of irregular mucosal surfaces by magnifying narrow-band imaging (ME-NBI), which offers high-resolution and detailed images of the esophagus. The overall boundary detection in diagnosing Lugol’s chromoendoscopy (LCE) images has been improved. This constitutes an effective and promising strategy for the precise detection of gastrointestinal lesions. The main contribution are as follows:

- Our proposed three-way fusion approach incorporates both macro and micro views, where macroscopic image provides overall boundaries of the esophagus, and micro image focuses on specific details. This integrated evaluation significantly improves the diagnostic accuracy. At patient level, our method shows that 43% of the patients who did not meet the 95% mean Average Precision (mAP) threshold should reach the standard threshold.
- The dual annotation in our approach effectively is capable of differentiating indistinct regions from non-lesion areas in ME-NBI and accurately capturing small lesions, which are the principle obstacle impeding precision in microscopic examination of ME-NBI.

## Methods

### 2.1 Three-way fusion method

From micro perspectives, ME-NBI imaging magnified lesions 80-110 times and presents several advantages in terms of enhancing diagnostic accuracy, lesion visibility, and image quality consistency. Additionally, it also provides better visualization of microvascular patterns and tissue structures, which aids physicians in making more precise assessments of lesion boundaries.[5] However, traditional ME-NBI CAD technology has yet to provide accurate treatment boundaries effectively. This is primarily due to the unevenness of lesions, resulting in distinct information on the main plane, while indistinct on side planes made the boundary ambiguous. Moreover, the focus of the image may vary, creating shadows in the image, and image reconstruction may result in blurriness due to motion artifacts, image processing, scatter radiation, and equipment issues. The final treatment boundary required in clinical practice comprises the distinct as well as indistinct area of the lesion. The indistinct area lies between the distinct lesion area and the non-lesion area, with ambiguous features, making the treatment boundary hard to determine. Additionally, shadows and blurriness can directly interfere with the treatment boundary determination.

To address these problems, we propose two annotation categories to make enhancements. 1) distinct boundary category (DBC): includes only the distinct portions of the lesion and excludes areas of indistinctness, shadowing, and blurriness. 2) comprehensive boundary category (CBC): includes both distinct and indistinct areas of the lesion while excluding focus and shadow interference areas. Physicians determine this boundary after reviewing all ME-NBI images of a patient comprehensively.

As a single ME-NBI image may not cover all areas of a lesion due to its magnification, physicians must review multiple images to diagnose the overall lesion region. Different images may capture the same region from different angles, resulting in indistinct areas being displayed as distinct in some images. Therefore, our dual annotation categories offer advantages over traditional CAD methods that only predict the CBC. Physicians can compare the prediction results between DBC and CBC across all related ME-NBI images, thus alleviating the problem of low reliability caused by indistinct areas’ unclear features. This dramatically improves the accuracy of treatment boundary prediction in clinical practice.

In contrast to traditional CAD technology, from macro perspectives, we introduced a novel technique called CAD-assisted LCE imaging, which provides precise treatment boundary visualization from a macroscopic perspective. Compared to ME-NBI imaging, LCE imaging has distinct advantages, including non-magnification, complete lesion boundary display, low equipment requirements, low medical costs, and a wide range of applications. Endoscopists have found LCE imaging to enhance the visualization of certain lesions, making it easier to identify areas of abnormal tissue. Additionally, LCE imaging can aid in the real-time diagnosis of neoplastic changes during endoscopy, potentially reducing the need for biopsy sampling. Notably, LCE imaging has demonstrated high sensitivity (>95%) in detecting EEC. [6]

### 2.2 Algorithm design

To validate the effectiveness of our three-way fusion method in improving EC lesion prediction accuracy and enhancing work productivity for clinical practitioners, we implemented eight different deep learning frameworks independently for each category. We developed diverse modules to address the challenges in identifying EC lesions. For instance, the lesion size demonstrates a range of scales, and different magnification levels in ME-NBI can exacerbate this challenge, resulting in the lesions exhibiting multiple scales. To address this, we introduced Deformable Convolutional Networks (DCN) [14] modules that adaptively scale the lesions.

Moreover, we encountered small-size lesions and boundary blurring. To tackle these problems, we incorporated various head structures, namely Cascade RoI Head [15], MaskScoring RoI Head [19], and Hybrid Task Cascade [17], which eliminated false detections and missed detections and amplified the segmentation’s precision and robustness. Additionally, we employed InstaBoost [13] to augment the model’s generalization ability. Besides, we choose Resnet101 and ResNeXt101 as our backbone and Feature Pyramid Network(FPN) as our neck.

### 2.3 Experimental dataset

We sourced data from patients diagnosed with EC between January 2010 and January 2020 at the First Affiliated Hospital of Shandong First Medical University. Our focus was on endoscopic images of patients diagnosed with high-grade dysplasia or T1 stage adenocarcinoma, with endoscopies performed using Olympus series upper endoscopes (260 HQ and 290 HQ; Olympus, Tokyo, Japan). We collected a total of 3500 images from 800 patients, including images of abnormal hyperplasia confirmed by EC histology. The histological evaluations were performed by four experienced gastrointestinal pathologists, and the endoscopy images were annotated by two endoscopists with over fifteen years of experience in surgical endoscopy. The criteria for defining non-abnormal EC images and annotation methodology were described in the appendix.

### 2.3 Evaluation methods

To measure the overall performance of the segmentation model, the Mean Average Precision (mAP) is used. mAP combines precision and recall to assess the model’s ability to detect and localize objects accurately. It is a widely used metric in evaluating object detection and segmentation algorithms.

For lesion detection at the pixel level, the average Intersection over Union (IoU) is provided to evaluate the quality of predicted bounding boxes or segmentation masks compared to the ground truth annotations. IoU quantifies the overlap between the predicted and ground truth regions, indicating the accuracy and precision of the predictions.

ARR (Average Recall Rate) measures the algorithm’s ability to accurately detect true positive objects.

It calculates the average ratio of the intersection area between the predicted and ground truth objects to the area of the ground truth objects. APR (Average Precision Rate) evaluates the precision of the algorithm in object detection. It calculates the average ratio of the intersection area between the predicted and ground truth objects to the area of the predicted objects. Both ARR and APR are valuable metrics for evaluating the performance of segmentation models, as they provide insights into the algorithm’s accuracy and reliability in detecting objects.

These metrics are evaluated on both bounding box and polygon levels, and the results of 24 experiments are reported at a 0.5 IoU level. Additional details and explanations of the evaluation metrics can be found in the Appendix.

## Results

### 3.1 Overall results

The obtained results demonstrate that the mAP for CBC is nearly 0.91, and the average IoU is 0.88, surpassing the mAP of 0.84 and the average IoU of 0.80 for DBC. This is consistent with our theoretical expectations, as CBC corresponds to the actual treatment boundary, while DBC only encompasses the distinct lesion area and improves the clinical accuracy of CBC. This suggests that in practical clinical settings, the accuracy of the treatment boundary is better than the performance of CBC. From a deep learning perspective, the superior performance of CBC over DBC implies that differentiating between non-lesion areas and indistinct lesion areas is easier than distinguishing indistinct and distinct lesion areas.

Using LCE category prediction effectively improved the precision of the clinical treatment boundary by providing a comprehensive view of the entire lesion. The evaluation of LCE using mAP and average IoU resulted in values of 0.86 and 0.84, respectively. It is important to note that these metrics are lower than CBC due to the more detailed depiction of the lesion provided by ME-NBI magnification. However, LCE poses a more challenging task than CBC as it covers the entire lesion boundary.

By comparing different frameworks, the hybrid task cascade ResNeXt101 with DCN was identified as the optimal framework across all categories, outperforming the others. Table 3 presents the Precision, Recall, and F1 Scores for all frameworks across all categories at both the bounding box and polygon levels. All eight frameworks performed well at the bounding box level, achieving an F1 score of more than 80%. Notably, using Cascade RoI Head for the region of interest selection resulted in better performance, primarily by reducing false positives. This approach also eliminates false detections and missed detections, further improving the model’s robustness. Moreover, all frameworks achieved higher Recall scores than Precision scores, which is desirable for clinical applications. A better Recall model would ensure a lower missing detection rate, minimizing the omission and negligence of EEC cases, which is critical for this application.

As shown in Table 1 and Table 2, the bounding box level outperformed the polygon level, as expected. Predicting polygons involves boundaries, making it more challenging to predict accurately. However, the performance of pixel-level polygons is more reliable, as it considers the actual area of the tumor lesion object and excludes any background noise interference.

**Table 1.** Image level performance based on Bounding Box.

| Distinct Boundary Category (DBC) on Bounding Box |  |  |  |  |
| --- | --- | --- | --- | --- |
| Model | avgIoU | APR | ARR | mAP |
| cascade_mask_rcnn+ResNet101+FPN+instaboost | 0.8180 | 0.8955 | 0.9119 | 0.8414 |
| cascade_mask_rcnn+ResNeXt101+FPN+DCN | 0.8087 | 0.8933 | 0.9041 | 0.8356 |
| cascade_mask_rcnn+ResNet101+FPN | 0.8044 | 0.8718 | 0.9207 | 0.7863 |
| mask_scoring_rcnn+ResNeXt101+FPN | 0.7926 | 0.8780 | 0.9018 | 0.8273 |
| hybrid_task_cascade+ResNeXt101+FPN | 0.8141 | 0.8874 | 0.9149 | 0.8127 |
| hybrid_task_cascade+ResNeXt101+FPN+DCN | 0.8233 | 0.9060 | 0.9081 | 0.8511 |
| mask_rcnn+ResNet101+FPN | 0.7965 | 0.8798 | 0.9020 | 0.8487 |
| hybrid_task_cascade+ResNeXt101+FPN+instaboost | 0.8202 | 0.8987 | 0.9124 | 0.8455 |
| Comprehensive Boundary Category (CBC) on Bounding Box |  |  |  |  |
| Model | avgIoU | APR | ARR | mAP |
| cascade_mask_rcnn+ResNet101+FPN+instaboost | 0.8866 | 0.9256 | 0.9570 | 0.9191 |
| cascade_mask_rcnn+ResNeXt101+FPN+DCN | 0.8773 | 0.9285 | 0.9432 | 0.9158 |
| cascade_mask_rcnn+ResNet101+FPN | 0.8795 | 0.9238 | 0.9518 | 0.9211 |
| mask_scoring_rcnn+ResNeXt101+FPN | 0.8684 | 0.9276 | 0.9334 | 0.9117 |
| hybrid_task_cascade+ResNeXt101+FPN | 0.8835 | 0.9270 | 0.9514 | 0.9059 |
| hybrid_task_cascade+ResNeXt101+FPN+DCN | 0.8865 | 0.9386 | 0.9433 | 0.9236 |
| mask_rcnn+ResNet101+FPN | 0.8643 | 0.9192 | 0.9385 | 0.9189 |
| hybrid_task_cascade+ResNeXt101+FPN+instaboost | 0.8877 | 0.9371 | 0.9463 | 0.9364 |
| Lugol's chromoendoscopy (LCE) on Bounding Box |  |  |  |  |
| Model | avgIoU | APR | ARR | mAP |
| cascade_mask_rcnn+ResNet101+FPN+instaboost | 0.8417 | 0.9083 | 0.9250 | 0.8315 |
| cascade_mask_rcnn+ResNeXt101+FPN+DCN | 0.8170 | 0.9068 | 0.8988 | 0.8042 |
| cascade_mask_rcnn+ResNet101+FPN | 0.8390 | 0.9144 | 0.9167 | 0.8332 |
| mask_scoring_rcnn+ResNeXt101+FPN | 0.8035 | 0.9026 | 0.8883 | 0.8122 |
| hybrid_task_cascade+ResNeXt101+FPN | 0.8305 | 0.9061 | 0.9155 | 0.8319 |
| hybrid_task_cascade+ResNeXt101+FPN+DCN | 0.8320 | 0.9070 | 0.9173 | 0.8620 |
| mask_rcnn+ResNet101+FPN | 0.7997 | 0.8938 | 0.8920 | 0.8434 |
| hybrid_task_cascade+ResNeXt101+FPN+instaboost | 0.8332 | 0.8999 | 0.9237 | 0.8408 |

**Table 2.** Image level performance based on Polygon Level.

| Distinct Boundary Category (DBC) on Polygon level |  |  |  |  |
| --- | --- | --- | --- | --- |
| Model | avgIoU | APR | ARR | mAP |
| cascade_mask_rcnn+ResNet101+FPN+instaboost | 0.7704 | 0.8679 | 0.8845 | 0.7947 |
| cascade_mask_rcnn+ResNeXt101+FPN+DCN | 0.7634 | 0.8796 | 0.8639 | 0.7829 |
| cascade_mask_rcnn+ResNet101+FPN | 0.7429 | 0.8544 | 0.8638 | 0.7165 |
| mask_scoring_rcnn+ResNeXt101+FPN | 0.7440 | 0.8712 | 0.8489 | 0.7709 |
| hybrid_task_cascade+ResNeXt101+FPN | 0.7570 | 0.8681 | 0.8676 | 0.7606 |
| hybrid_task_cascade+ResNeXt101+FPN+DCN | 0.7778 | 0.8868 | 0.8747 | 0.8277 |
| mask_rcnn+ResNet101+FPN | 0.7666 | 0.8778 | 0.8673 | 0.7871 |
| hybrid_task_cascade+ResNeXt101+FPN+instaboost | 0.7775 | 0.8768 | 0.8849 | 0.7915 |
| Comprehensive Boundary Category (CBC) on Polygon level |  |  |  |  |
| Model | avgIoU | APR | ARR | mAP |
| cascade_mask_rcnn+ResNet101+FPN+instaboost | 0.8619 | 0.9136 | 0.9406 | 0.8753 |
| cascade_mask_rcnn+ResNeXt101+FPN+DCN | 0.8483 | 0.9177 | 0.9219 | 0.8926 |
| cascade_mask_rcnn+ResNet101+FPN | 0.8537 | 0.9141 | 0.9319 | 0.8935 |
| mask_scoring_rcnn+ResNeXt101+FPN | 0.8445 | 0.9208 | 0.9128 | 0.8792 |
| hybrid_task_cascade+ResNeXt101+FPN | 0.8559 | 0.9174 | 0.9310 | 0.8795 |
| hybrid_task_cascade+ResNeXt101+FPN+DCN | 0.8511 | 0.9245 | 0.9181 | 0.9079 |
| mask_rcnn+ResNet101+FPN | 0.8439 | 0.9163 | 0.9172 | 0.8848 |
| hybrid_task_cascade+ResNeXt101+FPN+instaboost | 0.8562 | 0.9246 | 0.9243 | 0.9087 |
| Lugol's chromoendoscopy (LCE) on Polygon level |  |  |  |  |
| Model | avgIoU | APR | ARR | mAP |
| cascade_mask_rcnn+ResNet101+FPN+instaboost | 0.7899 | 0.8892 | 0.8800 | 0.8423 |
| cascade_mask_rcnn+ResNeXt101+FPN+DCN | 0.7753 | 0.8866 | 0.8657 | 0.7960 |
| cascade_mask_rcnn+ResNet101+FPN | 0.7867 | 0.8990 | 0.8679 | 0.8445 |
| mask_scoring_rcnn+ResNeXt101+FPN | 0.7753 | 0.8933 | 0.8602 | 0.8325 |
| hybrid_task_cascade+ResNeXt101+FPN | 0.7897 | 0.8967 | 0.8738 | 0.8403 |
| hybrid_task_cascade+ResNeXt101+FPN+DCN | 0.7933 | 0.8984 | 0.8756 | 0.8501 |
| mask_rcnn+ResNet101+FPN | 0.7744 | 0.8944 | 0.8571 | 0.8545 |
| hybrid_task_cascade+ResNeXt101+FPN+instaboost | 0.7914 | 0.8889 | 0.8832 | 0.8295 |

### 3.2 Image level

ME-NBI imaging can be classified into two categories: Whole Lesion Coverage Imaging (WLCI) and Incomplete Lesion Coverage Imaging (ILCI). In WLCI, the entire image is encompassed by CBC, while in ILCI, only a portion of the image is covered by CBC.

#### 3.2.1 Whole Lesion Coverage Imaging

In our study, we found that 36% of all ME-NBI images fall under the category of WLCI. In such cases, the image only captures a partial area of the lesion, and the clinical treatment boundary lies outside the image. Our analysis of the annotations showed that the area of indistinct regions in WLCI is not insignificant. In some cases, the edges mainly consist of indistinct regions, as shown in (Figure 2a).

**Figure 1.**
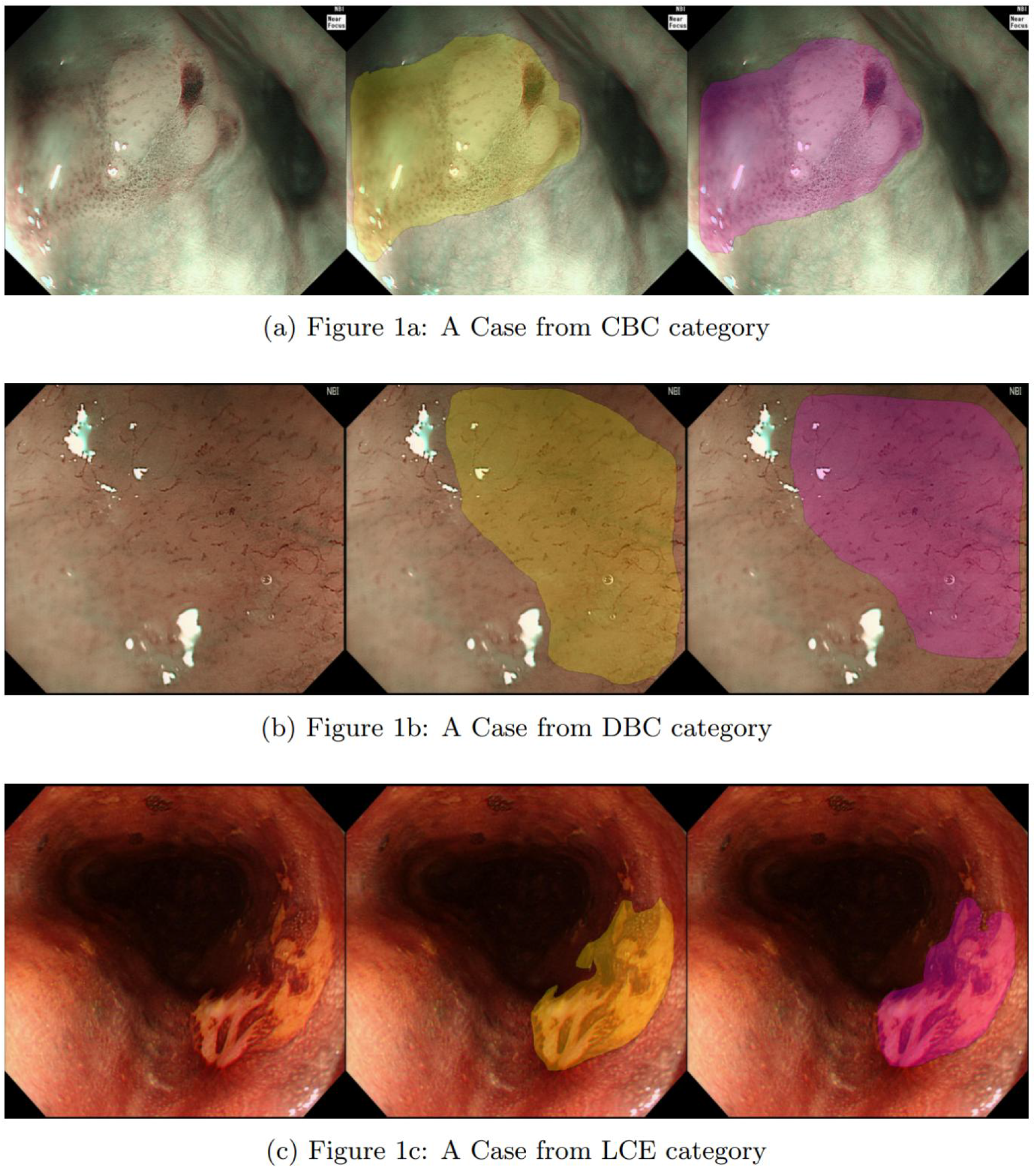
shows predicted examples for categories CBC, DBC, and LCE, arranged from top to bottom, respectively. Each subfigure presents the original image, annotated image, and predicted image from left to right. The predicted lesions were obtained using cascade_mask_rcnn_r101_fpn_instaboost framework for CBC category (Figure 1a), cascade_mask_rcnn_x101_324d_fpn_dconv_c3-c5 framework for DBC category (Figure 1b), and cascade_mask_rcnn_x101_324d_fpn_dconv_c3-c5 framework for LCE category (Figure 1c).

**Figure 2a.**
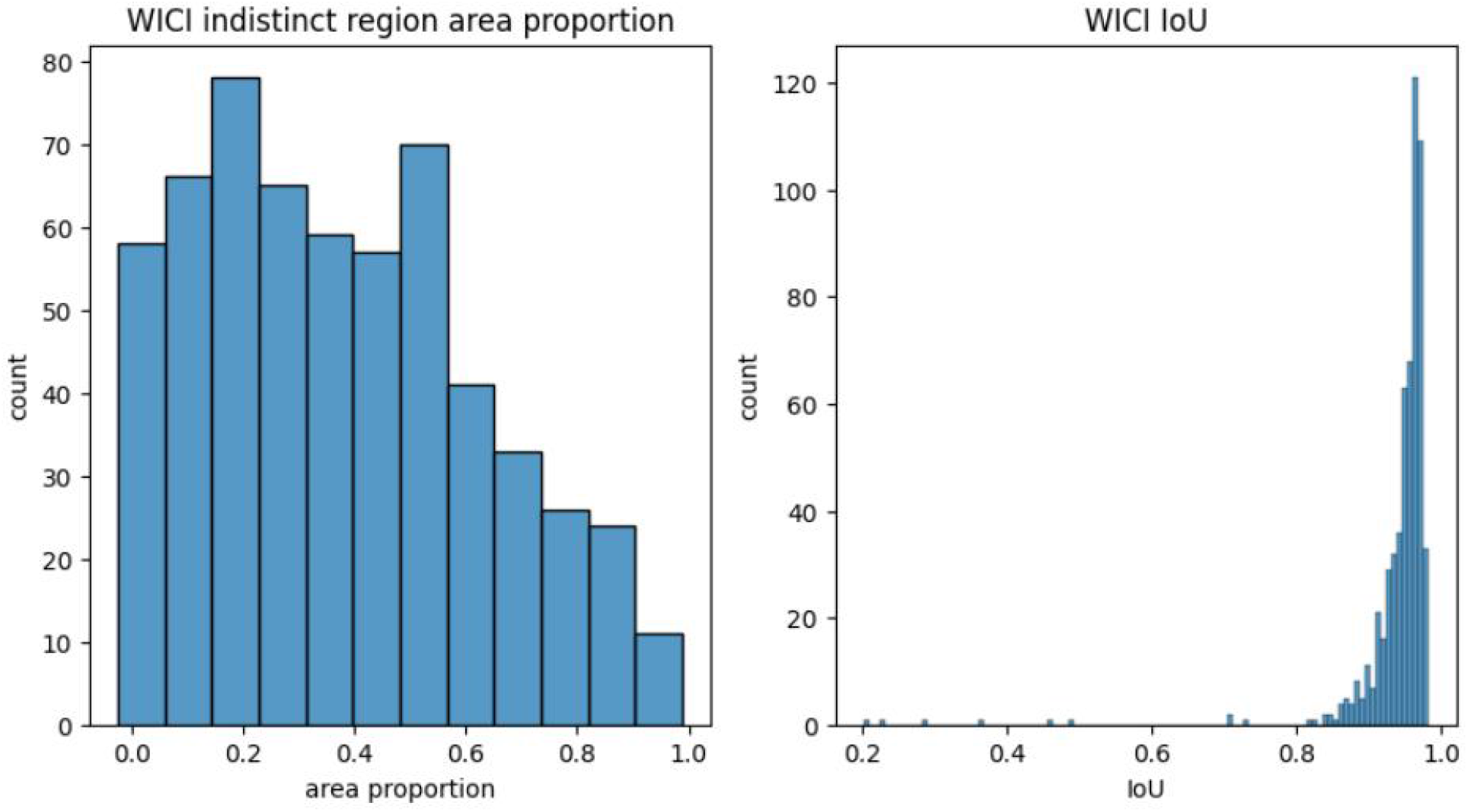
shows WLCI indistinct region area proportion, the area occupied by indistinct regions in WICI is non-negligible and should not be ignored. Figure 2b shows IoU histogram on the object level, with an average greater than 0.95.

Physicians differentiate indistinct regions as either lesions or non-lesions only after careful consideration, comparison, and discussion during the image annotation process. However, in actual clinical practice, physicians may face challenges in quickly distinguishing between indistinct lesion areas and non-lesion areas and eliminating shadows and blurriness. Our CAD system’s performance in WLCI on CBC shows a nearly 1.00 mAP and an average IoU greater than 0.95 (Figure 2b). In other words, our CAD system can assist physicians in making judgments on indistinct regions in clinical practice for WLCI promptly.

#### 3.2.2 Incomplete Lesion Coverage Imaging

ILCI can be further subdivided into two groups based on the proportion of indistinct areas in the image. The first group comprises images that do not feature any indistinct regions, leading to the CBC region being the same as the DBC region. The second group, however, contains indistinct regions, resulting in the CBC region being more extensive than the DBC region, as depicted in (Figure 3a). In both categories, physicians must determine the clinical treatment boundary.

**Figure 3a.**
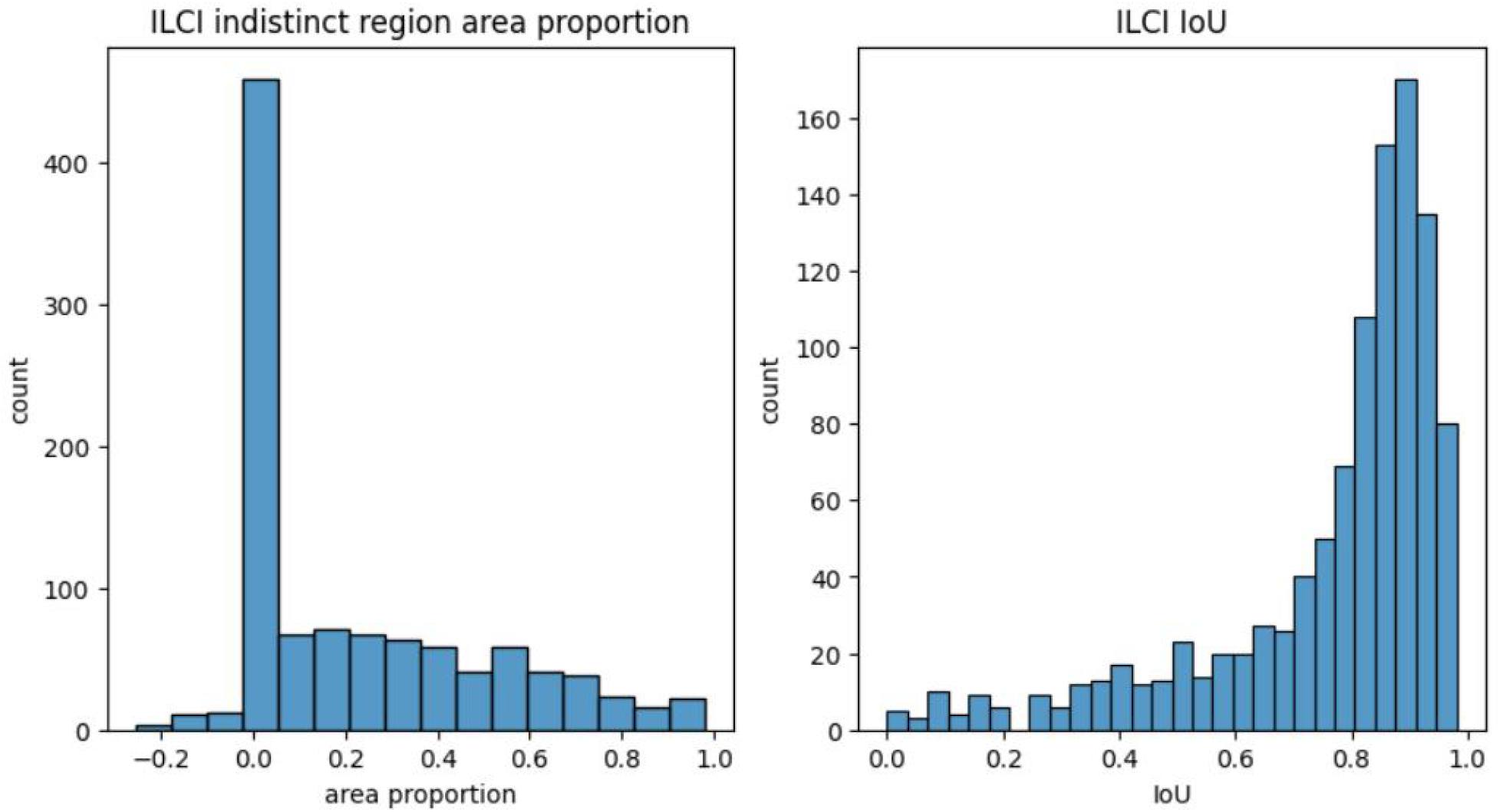
shows ILCI indistinct region area proportion, the area occupied by indistinct regions in ILCI is significantly smaller than that of WLCI regions. Figure 3b shows IoU histogram on the object level still demonstrates good performance.

Our experimental findings reveal that ILCI achieves an mAP of 0.93 in CBC, and the distribution of IoU values is illustrated in Figure 3b. Despite ILCI achieving a high mAP score, indicating excellent performance, it still performs slightly worse than WLCI in CBC detection. This observation suggests that differentiating between the indistinct lesion area and the non-lesion area is a challenging task, and ILCI tends to predict a larger region than the ground truth region. Specifically, ILCI exhibits higher sensitivity but lower specificity, making it more prone to falsely predicting non-lesion indistinct areas as lesions and unable to resist shadow and blur interference.

To address these issues, our CAD system incorporates DBC prediction to mitigate the problems. For instance, in first image, the ILCI predicted an indistinct non-lesion area as a lesion in the CBC category, while the DBC prediction in second image correctly excluded the same area. Our study highlights the importance of effective dual annotation methods to aid physicians in accurately diagnosing and treating lesions, compared to traditional CAD systems that only offer predictions on the CBC category.

The correlation between the proportion of indistinct areas and the IoU is only 0.1. The t-test results for the proportion of indistinct areas between the IoU>0.5 and IoU<0.5 buckets are insignificant. This suggests that the IoU is not directly related to the proportion of indistinct areas but instead to the distinctiveness of the lesion boundary.

DBC outperforms CBC, with an average IoU of 0.79 compared to 0.74 for 424 images without indistinct regions. Smaller lesions are more challenging to predict, but DBC performs significantly better than CBC for smaller-size lesions, with an average IoU of 0.74 compared to 0.65 for CBC. Our findings suggest that DBC can better assist physicians in determining the treatment boundary, especially when all lesions are in a distinct state and when the lesion area is small, compared to only CBC annotation in traditional CAD systems.

Figure 4 shows a small lesion with distinct regions, where (a), (b), and (c) represent the annotated, CBC-predicted, and DBC-predicted areas, respectively. Our analysis reveals that CBC’s prediction result is interfered with by the side plane indistinct regions and focus-induced glare, resulting in falsely predicted areas. However, the DBC prediction result in (c) is highly accurate, with an IoU of up to 98%. These results demonstrate the effectiveness of our innovative DBC category in accurately predicting lesion boundaries, especially in cases where all lesions are in a distinct state.

**Figure 4a,4b,4c.**
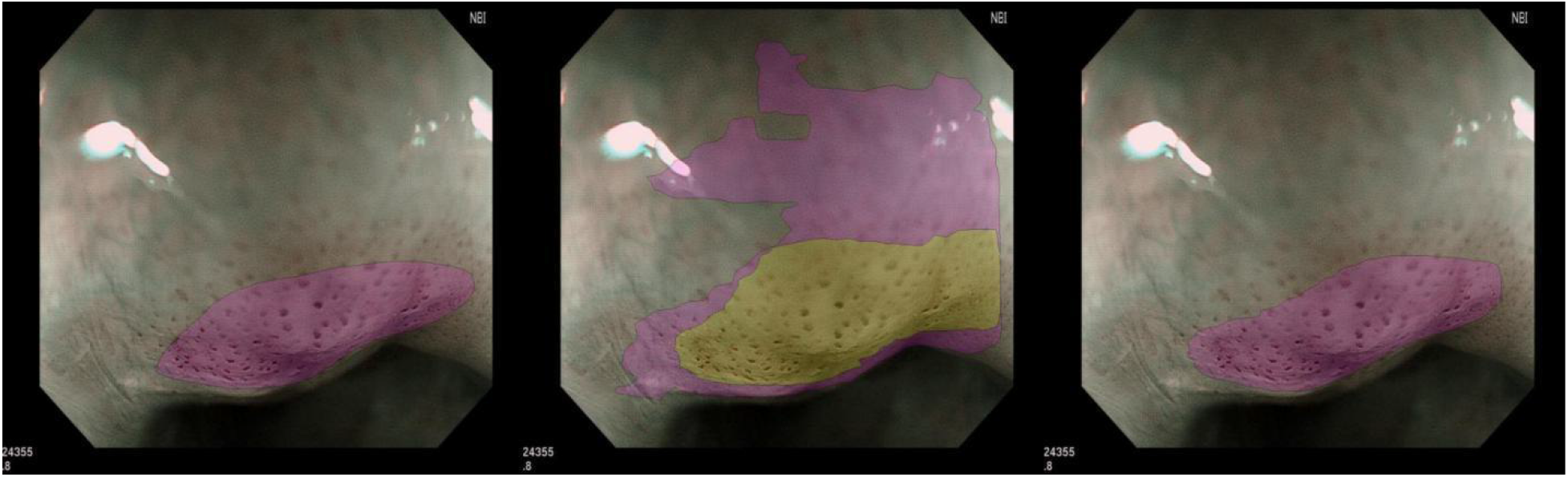
An example with Annotated region, CBC predicted region, and DBC predicted region from left to right.

### 3.3. Patient level

On average, a patient may have 4-5 ME-NBI images, with some patients as many as 40. At the patient level, the mAP in CBC can reach up to 0.95. For patients below 0.95, after our CAD system assists in the DBC category, there are at least 43% of patients have mAP higher than 0.95, indicating that our CAD system’s high performance is not limited to the high diagnostic rate of CBC but also extends to assisting patients who do not meet the standard line through DBC. As a result, the performance of the clinical treatment boundary at the patient level is also superior to that of CBC and also the traditional single CBC annotation CAD.

### 3.4 Lugol’s chromoendoscopy

Compared to ME-NBI images, LCE images exhibit higher sensitivity in lesion boundaries but lower specificity. Furthermore, certain non-tumorous regions with inflammation can also display shallow staining areas, leading to potential overestimation of the predicted area compared to the ground truth. However, in our CAD system, the APR and ARR are generally equal, indicating that the predicted area is similar to the ground truth area. Additionally, the IoU performance is not directly influenced by the staining chromatics of the lesion or the color difference between the lesion and non-lesion regions. This is because we conducted color normalization on LCE images during model training, providing the model with solid robustness in distinguishing between lesion and non-lesion regions under different staining effects and identifying diverse chromatic differences caused by staining. Furthermore, akin to ME-NBI, the IoU performance of LCE is not related to the size of the lesion itself but rather to the disparity in the image at the junction between the lesion and non-lesion regions.

### 3.5 Causes of FN, FP

In most cases where ME-NBI images are falsely predicted, the model erroneously identifies non-lesion areas as lesions, particularly in CBC. Two common factors that contribute to such outcomes are indistinct areas on the side plane that make it challenging for the model to distinguish accurately, as seen in (Figure 4b), and interference caused by imaging flashes and shadows, as depicted in (Figures 5), which respectively display the CBC labeled area and predicted area. The glaring point in the upper left corner and the shadow area in the upper right corner both cause interference with the model’s prediction. Additionally, the DBC IoU in this example is higher than 0.90, which can assist physicians.

**Figure 5a,5b.**
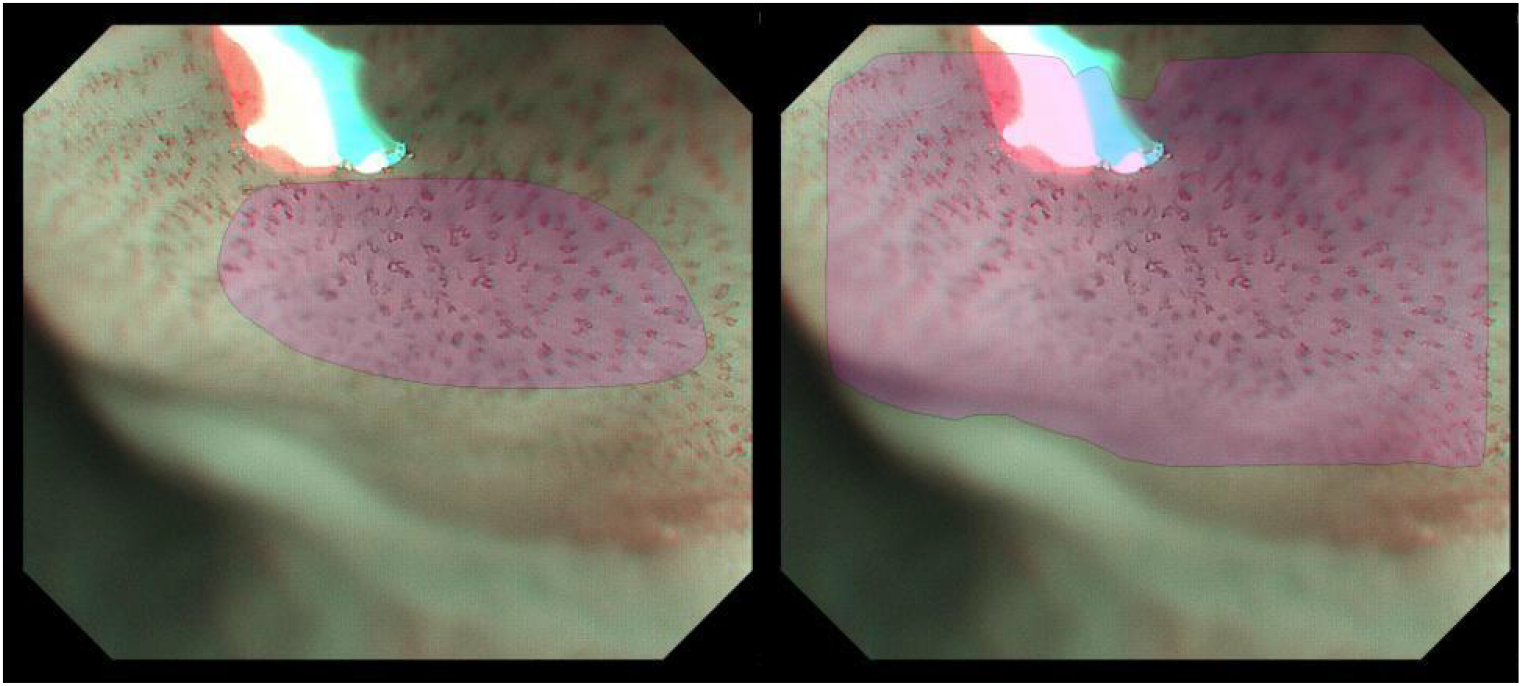
Interference caused by imaging flashes and shadows. CBC label and predicted region shows from left to right.

## Discussion

Our study demonstrates the effectiveness of a novel three-way fusion CAD system for detecting EC. Our findings indicate that using the DBC category significantly improves the accuracy of lesion detection and boundary delineation, and the LCE category optimizes the accuracy from an overall perspective. Moreover, our three-way fusion CAD system achieved promising results for both the ME-NBI and LCE categories, surpassing the performance of traditional CAD systems.

The superiority of our CAD system can be attributed to several factors. First, we used a hybrid task cascade framework with DCN, which proved to be effective in accurately predicting the lesion region with different scales. Second, we incorporated LCE category prediction, which enabled a comprehensive view of the entire lesion. Third, the use of DBC prediction significantly reduced false detections and missed detections, improving the model’s robustness and emphasizing the importance of effective dual annotation methods to aid physicians in accurately diagnosing and treating lesions.

Our study also revealed some challenges and limitations. In ME-NBI imaging, the presence of indistinct regions can interfere with accurate lesion detection and boundary delineation. Furthermore, smaller lesions are more difficult to predict accurately. In LCE imaging, non-tumorous regions with inflammation can also display shallow staining areas, potentially leading to overestimating the predicted area compared to the ground truth. However, our CAD system demonstrated robustness in distinguishing between lesion and non-lesion regions under different staining effects and chromatic differences.

Future studies can further improve the performance of CAD systems for EEC detection by incorporating other imaging modalities. Additionally, using multi-modality and ensemble learning can improve the robustness and generalizability of CAD systems. Finally, our study highlights the importance of continuous data collection and analysis to optimize CAD systems for clinical applications and improve patient outcomes.

## Conclusion

In this paper, we proposed a novel three-way fusion CAD system for detecting early EC using ME-NBI and LCE imaging. Our system outperforms traditional CAD methods that only predict the CBC by incorporating the DBC category. We conducted 24 experiments on two ME-NBI categories and one LCE category, demonstrating promising results across all categories. Our best results for the CBC category were achieved using the hybrid task cascade ResNeXt101 with DCN, with an mAP of 0.9236, APR of 0.907, and ARR of 0.9173 at a 0.5 threshold IoU level. We also achieved high precision and recall of 93.98% and 93.05%, respectively. The LCE category showed the best results using the hybrid task cascade framework, with an mAP of 0.8620, APR of 0.9386, and ARR of 0.9433, along with 82.89% precision and 88.32% recall. Our three-way fusion method demonstrated excellent diagnostic performance with accurate treatment boundaries on both image and patient levels. These results confirm the feasibility of deep learning for detecting EC in both ME-NBI and LCE.

Overall, our results highlight the importance of incorporating the DBC category in CAD systems, which has not been previously explored in the literature. This approach can assist physicians in accurately diagnosing and treating lesions, especially in cases where all lesions are in a distinct state or incomplete lesion coverage imaging. Our study also emphasizes the effectiveness of utilizing deep learning techniques for detecting EC, showcasing promising performance across different imaging categories. Future work can involve collecting and analyzing patient data from a broader area to achieve real-time performance for clinical applications.

## Data Availability

Due to the nature of this research, participants of this study did not agree for their data to be shared publicly, so supporting data is not available.

## Appendix Annotation Criteria

In the course of processing the experimental dataset, we meticulously followed a set of plans and procedures that encompassed defining the non-abnormity of EEC images and performing annotations. The corresponding details are presented below.

To classify an image as non-abnormity, it had to meet all of the following three criteria: 1) The patient had no prior history of esophageal lesions detected during examination. 2) Confirmation was obtained from endoscopists with over a decade of experience in endoscopy. 3) Unanimous agreement was reached on the non-abnormity status.

For the annotation of abnormity images, we proposed an annotation methodology and adhered strictly to the following steps to ensure consistency and professionalism: 1) The first endoscopist reviewed the gastroscopy image and marked the lesion as a polygon. 2) The second endoscopist examined the annotation results and added notes as necessary. 3) If there were no disagreements, the annotation was considered complete. If a disagreement arose, a third endoscopist was consulted, and the three endoscopists collaborated to arrive at the final decision.

As a single ME-NBI image may not cover all areas of a lesion due to its magnification, physicians must review multiple images to diagnose the overall lesion region. Commonly, there are 4 to 5, up to 40 ME-NBI images per patient.

### Evaluation Metrics

Details, explanations and equations of the evaluation metrics:

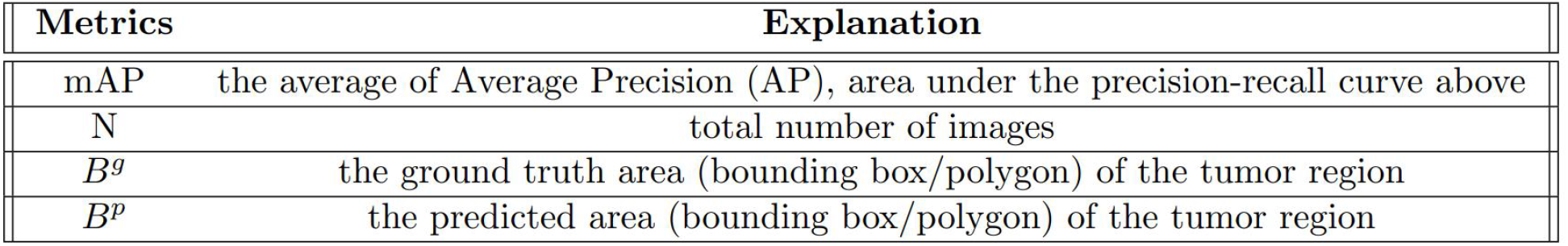

| Metrics | Explanation |
| --- | --- |
| mAP | the average of Average Precision (AP), area under the precision-recall curve above |
| N | total number of images |
| $B^g$ | the ground truth area (bounding box/polygon) of the tumor region |
| $B^p$ | the predicted area (bounding box/polygon) of the tumor region |

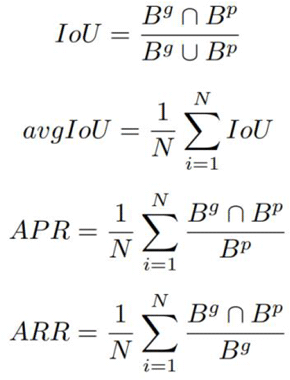

